# Long-term outcomes and GVHD in patients receiving hematopoietic cell transplants abroad: A 14-year UAE multi-center cohort

**DOI:** 10.1101/2025.11.15.25340313

**Authors:** Naveed Syed, Mohamed Abuhaleeqa, Fatema Mohammed Al Kaabi, Naser Alzein, Zainul Aabideen, Aydah M. Al-Awadhi, Arif Alam, Azmat Ali Khan, Imrana Afrooz, Farooq Ahmed Mir, Riad Alhasan, Moussab Damlaj, Osama Taher Anaam, Manar Hassan Mohamed, Gehad ElGhazali, Kayane Mheidly, Shahrukh Hashmi

## Abstract

**Background:** Hematopoietic cell transplant (HCT) outcomes are well characterized in clinical trials and international registries, but data remain scarce for patients who undergo transplantation abroad and return home for follow-up. While transplant centers typically report procedural details and early mortality for this cohort, long-term survival and chronic graft versus host disease (GVHD) outcomes are seldom captured, underscoring the need for comprehensive reporting.

**Methods:** We conducted a retrospective, multi-center study of all HCT recipients followed at all the three tertiary hospitals in Abu Dhabi, UAE, between 2009 and 2023. Patient demographics, transplant characteristics, overall survival (OS), and GVHD incidence were extracted from medical records. Kaplan–Meier OS estimates were calculated for diseases with ≥20 cases; crude survival was reported for rarer indications.

**Results:** A total of 454 HCTs were analyzed: 350 (77.1%) allogeneic and 104 (22.9%) autologous. Adults undergoing allogeneic HCT were relatively young (median age 35.3 years), reflecting referral practices and the UAE’s demographic profile. An unexpected female predominance was observed in pediatric malignancies and immune deficiencies. Matched unrelated donor (MUD) transplants were rare, and the trend remained low over time, underscoring reliance on matched related donor (MRD) and mismatched related donor (MMRD). The incidence of GVHD was high, approaching the upper range of international reports: 69.2% in adults and 45.7% in children. In adults with AML, acute GVHD was associated with an inferior 5-year OS (67.9% vs. 86.2%, p=0.036), whereas chronic GVHD did not significantly impact outcomes. In pediatric AML, both acute and chronic GVHD were linked to poorer survival (<50% at 5 years), while outcomes for B/T-ALL, thalassemia, and SCID exceeded 85–90% at 5 years regardless of GVHD status.

**Conclusions:** HCT outcomes in this transplant tourism cohort were generally comparable to international benchmarks, yet notable for younger adult recipients, predominance of allogeneic over autologous procedures, limited use of MUDs, and a high incidence of GVHD. These observations highlight the urgent need to establish sustainable local transplant capacity, build donor registries, and implement aggressive GVHD prevention and treatment practices to improve outcomes in the UAE.

## 1 Introduction

Hematopoietic cell transplantation (HCT) remains the only curative option for a broad spectrum of benign and malignant hematological disorders[1][2][3]. However, graft-versus-host disease (GVHD) remains a leading cause of transplant-related morbidity and mortality[4][5][6]. Transplant outcomes are often assessed by overall survival (OS)—the gold-standard endpoint and, specifically for allogeneic HCT, by the incidence and severity of acute and chronic GVHD along with the transplant related mortality (TRM).

The global landscape of HCT is well documented by the Worldwide Network for Blood and Marrow Transplantation (WBMT)[9]. Global HCT activity has more than doubled over the past decade, reflecting broader access and evolving donor strategies, particularly the expanding use of haploidentical donors. Despite these advances and robust international reporting, substantial gaps remain in reporting between regions. In the United Arab Emirates (UAE): historically, most patients travelled abroad for receipt of HCT, and while the recent establishment of local programs has begun to reduce this reliance, published outcome data for those transplanted internationally but followed in home country are lacking[10][11].

Preliminary insights were provided by our pilot study, which reported a high prevalence of GVHD (62.2%) among patients transplanted abroad, despite a predominantly young cohort and a high proportion of HLA-identical related donors [12]. These findings suggest that factors inherent to the “transplant tourism” model, such as fragmented care and protocol heterogeneity, may adversely influence outcomes. However, that study was limited to a single-center experience with a modest cohort and short follow-up.

To address these gaps, we conducted a comprehensive, multi-center analysis of potentially all HCT patients followed in the UAE capital over a 14-year period. The objectives were to: (1) describe allogeneic and autologous transplant activity, including indications across adult and pediatric populations; (2) evaluate long-term OS by key disease groups; and (3) assess the incidence of acute and chronic GVHD in a larger, more representative cohort. This study establishes essential benchmarks for HCT outcomes in this unique cohort and provides critical evidence to promote the development of sustainable local transplant programs and national health policy.

## 2 Methods

### 2.1 Patient identification and data collection

Following approval from the Institutional Review Boards of all participating centers (SSMC REC-444, Date: 11/30/2022), patient data were retrospectively extracted from the electronic medical records (EMR) of all the three tertiary centers in Abu Dhabi: Sheikh Shakhbout Medical City, Tawam Hospital, and Sheikh Khalifa Medical City. Data collection was carried out by a team of physicians, research coordinators, and rotating medical students. A keyword search was performed using terms such as *bone marrow transplant*, *stem cell transplant*, *graft-versus-host disease (acute or chronic)*, and *hematopoietic stem cell transplant*. Data were collected using a standardized electronic case report form (eCRF). To ensure consistency, all data abstractors received training on the protocol, and ambiguous cases were reviewed by a senior study investigator (N.S.).

Between 2009 and 2023, a total of 598 patients were identified. Of these, 454 patients were included in the analysis. Exclusions (n = 144) were primarily due to fewer than three follow-up clinic visits, insufficient data, patients who underwent transplantation but did not return for follow-up, or patients who received solid organ transplants. Individuals transplanted prior to 2009 were also excluded owing to incomplete transplant-related data.

### 2.2 Patient classification and definitions

Patients aged ≥18 years at the time of transplantation were classified as adults. For malignant disorders, the date of diagnosis was defined by the bone marrow biopsy report. For non-malignant conditions (e.g., inherited bone marrow failure syndromes or immune deficiencies), the date of first clinical documentation of the disorder was taken as the diagnosis date. Patients who underwent a second transplant were treated as independent cases, given the changes in donor characteristics, disease status, or primary diagnosis.

### 2.3 Transplant characteristics and GVHD Definitions

Stem cell source (bone marrow, peripheral blood, or cord blood) was documented. Conditioning regimens were abstracted by drug names due to incomplete recording of conditioning intensity. Human leukocyte antigen (HLA) compatibility was defined as follows:

- **Full match**: 10/10, 8/8, or 6/6
- **Partial match**: 9/10
- **Mismatched**: all others

Graft-versus-host disease (GVHD) was categorized as acute or chronic based on physician documentation based on NIH GVHD consensus criteria, and alternatively with onset within 100 days post-transplant classified as acute GVHD and onset beyond 100 days classified as chronic GVHD in the case of missing NIH consensus GVHD criteria. As patients underwent transplantation at various centers worldwide, definitions and severity grading were inconsistent; therefore, we applied the ≤100 and >100-day cutoffs in accordance with NIH GVHD consensus criteria to ensure uniform classification in the latter group. Severity of GVHD, transplant conditioning regimens, and GVHD prophylaxis strategies were not reported owing to inconsistencies in documentation and limited detail available from physician records from international centers. We aimed to report only genuine findings and restrict analyses to datapoints that could be extracted with confidence and accuracy.

### 2.4 Study outcomes and survival Analysis

The final study population comprised 454 patients, of whom 230 (50.7%) were pediatric and 224(49.3%) were adult cases. Age classification was based on age at the time of transplant. Overall survival (OS) was defined as the time from transplant until death from any cause or last known follow-up. Cause-specific survival (transplant-related vs. non-transplant-related mortality) could not be reliably determined due to incomplete reporting. Kaplan-Meier survival curves were generated with confidence intervals were calculated for disease categories with ≥20 patients; for all other indications, crude survival rates were reported and details included in supplementary documents. Comparisons between groups were provided where sample sizes were permitted.

## 3 Results

### 3.1 Transplant activity and patient characteristics

#### 3.1.1 Transplant activity and indications

A total of 454 hematopoietic stem cell transplants (HCTs) were identified, comprising 350(77.1%) allogeneic and 104(22.9%) autologous transplants. The median age of adults undergoing allogenic HCT was 35.3 years (IQR, 26.6–49.1). As seen in **Table 1**. Transplant activity for malignant and non-malignant disorders (2009–2023), non-malignant disorders were the leading indication for allogeneic HCT (193, 42.5%), followed by lymphoid (156, 34.4%) and myeloid (93, 20.5%) malignancies, while solid tumors represented a minority (12, 2.6%). Within the non-malignant group, thalassemia (17.4%) and primary immune deficiencies such as severe combined immunodeficiency (SCID, 9.3%) were the most frequent indications. In the malignant cohort, acute myeloid leukemia (AML, 16.7%) was the major myeloid indication, whereas acute lymphoblastic leukemia (ALL, 14.1%) predominated in the lymphoid group. Among autologous transplants, plasma cell disorders (9.9%) were the most common.

**Table 1.** Transplant activity for malignant and non-malignant disorders (2009–2023)

| **Disease Category** | **Allogeneic, n (%)** | **Autologous, n(%)** | **Total, n (%)** |
| --- | --- | --- | --- |
| **Myeloid malignancies** | **79 (84.9)** | **14 (15.1)** | **93 (20.5)** |
| › Acute myeloid leukemia | 64 (84.2) | 12 (15.8) | 76 (16.7) |
| › Chronic myeloid leukemia | 2 (66.7) | 1 (33.3) | 3 (0.7) |
| › MPN/MDS/BPDCN | 13 (92.9) | 1 (7.1) | 14 (3.1) |
| **Lymphoid malignancies** | **78 (50)** | **78 (50)** | **156 (34.4)** |
| › Acute lymphoblastic leukemia | 63 (98.4) | 1 (1.6) | 64 (14.1) |
| › Chronic lymphocytic leukemia | 1 (100) | 0 | 1 (0.2) |
| › Plasma cell disorders (MM) | 4 (8.9) | 41 (91.1) | 45 (9.9) |
| › Hodgkin lymphoma | 4 (23.5) | 13 (76.5) | 17 (3.7) |
| › Non-Hodgkin lymphoma | 6 (20.7) | 23 (79.3) | 29 (6.4) |
| **Solid tumors** | **0** | **12 (100)** | **12 (2.6)** |
| › Neuroblastoma | 0 | 9 (100) | 9 (2.0) |
| › Medulloblastoma | 0 | 3 (100) | 3 (0.7) |
| **Non-malignant disorders** | **193 (100)** | **0** | **193 (42.5)** |
| › Thalassemia ($\alpha + \beta$) | 79 (100) | 0 | 79 (17.4) |
| › Sickle cell disease | 25 (100) | 0 | 25 (5.5) |
| › Bone marrow failure* | 31 (100) | 0 | 31(6.8) |
| › Primary immune deficiencies** | 42 (100) | 0 | 42 (9.3) |
| › Others*** | 16 (100) | 0 | 16 (3.5) |
| **Total** | **350 (77.1)** | **104 (22.9)** | **454** |

\*Bone marrow failure: Aplastic anemia, Fanconi anemia, dyskeratosis congenita, Paroxysmal nocturnal hemoglobinuria
\*\*PIDs: Severe combined immune deficiency, chronic granulomatous disease, Hyper IgM syndrome, common variable immune disorder
\*\*\*Others: Hemophagocytic lymphohistiocytosis (8), Sideroblastic anemia (1), Amyloidosis (1), Erythrogenic porphyria (1), Glanzman thrombasthenia (2), B mannosidosis (1), Langerhans cell histiocytosis (1)

As illustrated in **Supplemental Figure 1**, the annual number of allogeneic transplants steadily increased over the study period, reaching a peak in 2021 before declining thereafter. In contrast, autologous transplants remained lower in number, with a fluctuating trend and smaller peaks noted in 2017, 2019, and 2021.

#### 3.1.2 Age and gender distribution

As seen in **Table 2**, among allogeneic HCT recipients, 133 (36.6%) were adults and 230 (63.4%) were pediatric patients. In myeloid malignancies, the majority were adults (60, 65.2%) with a median age of 45.5 years (IQR, 33.1–51.8), while pediatric recipients accounted for 32 (34.8%) with a median age of 8.0 years (IQR, 3.5–13.9). Similarly, in lymphoid malignancies, 52 (65.8%) were adults (median age, 33.4 years), compared to 27 (34.2%) pediatric cases (median age, 6.6 years). In contrast, non-malignant disorders were predominant in the pediatric population (171, 89.1%) with a median age of 5.7 years (IQR, 3.0–10.3), whereas only 21 (10.9%) adults underwent HCT for these indications (median age, 27.0 years). Male recipients predominated across all three adult groups, while females were the majority among children transplanted for myeloid and lymphoid malignancies.

**Table 2.** Baseline characteristics of HCT recipients by disease group (2009–2023)

| Disease group | Adults |  |  | Pediatrics |  |  |
| --- | --- | --- | --- | --- | --- | --- |
|  | n (%) | Age, y (IQR) | Male, n (%) | n (%) | Age, y (IQR) | Male, n (%) |
| **Allo-HCT** |  |  |  |  |  |  |
| › Myeloid | 60 (65.2) | 45.5 (33.1–51.8) | 37 (61.6) | 32 (34.8) | 8.0 (3.5–13.9) | 15 (46.8) |
| › Lymphoid | 52 (65.8) | 33.4 (25.0–43.5) | 33 (63.5) | 27 (34.2) | 6.6 (4.1–12.7) | 12 (44.4) |
| › Non-malignant † | 21 (10.9) | 27.0 (19.7–30.9) | 11 (52.4) | 171 (89.1) | 5.7 (3.0–10.3) | 95 (55.5) |
|  | **133** |  |  | **230** |  |  |
| **Auto-HCT** |  |  |  |  |  |  |
| › Lymphoid | 74 (96.1) | 50.1 (40.3–57.4) | 45 (60.8) | 3 (3.9) | 14.6 (9.7–15.2) | 3 (100) |
| › Solid tumors | - | - | - | 12 (100) | 3.7 (3.3–6.0) | 6 (50.0) |
| › Others ^ | 2 | - | - | - | - | - |
|  | **76** |  |  | **15** |  |  |

Abbreviations: HCT, hematopoietic stem cell transplantation; IQR, interquartile range.
† Includes HLH, porphyria, Glanzmann thrombasthenia, $\beta$ -mannosidosis, Langerhans-cell histiocytosis, sideroblastic anemia.
^ Includes *amyloidosis* (1), *AML* (1)

As summarized in **Table 2**, autologous HCT was reported in adults (76, 83.5%), with pediatric cases accounting for only a small number of patients (15,16.5%). The majority of adult recipients had transplant for lymphoid malignancies (74, 96.1%), with a median age of 50.1 years (IQR, 40.3–57.4), and 60.8% were male. In contrast, pediatric autologous HCT was almost exclusively performed for solid tumors (12, 100%), with a median age of 3.7 years (IQR, 3.3–6.0), while a small proportion of pediatric cases (3, 3.9%) were lymphoid malignancies. Male predominance was noted among pediatric lymphoid cases (100%) but was less pronounced in solid tumors (50%). For detailed distribution of specific diagnoses and corresponding patient age and sex characteristics among both allogeneic and autologous HCT recipients, refer to **Supplemental Tables 1 and 2**.

#### 3.1.3 Donor type distribution and temporal trends

As detailed in **Supplemental Table 3**, donor type distribution varies across disease categories and age groups. Among allogeneic transplants, MRD were the predominant source in myeloid and lymphoid malignancies, while MMRD were most frequently utilized in non-malignant disorders, particularly for thalassemia and severe combined immunodeficiency (SCID). MUD was a smaller proportion, largely restricted to adult AML and lymphoid malignancies. Temporal trends are illustrated in **Figure 1**, demonstrating a steady rise in MMRD use from 2013 onwards, in parallel with a gradual decline in MUD transplants. MRD was the predominant donor source, followed by MMRD and MUD, as shown in **Supplemental Table 4**, with a similar distribution observed in both adult and pediatric cohorts.

**Figure 1.**
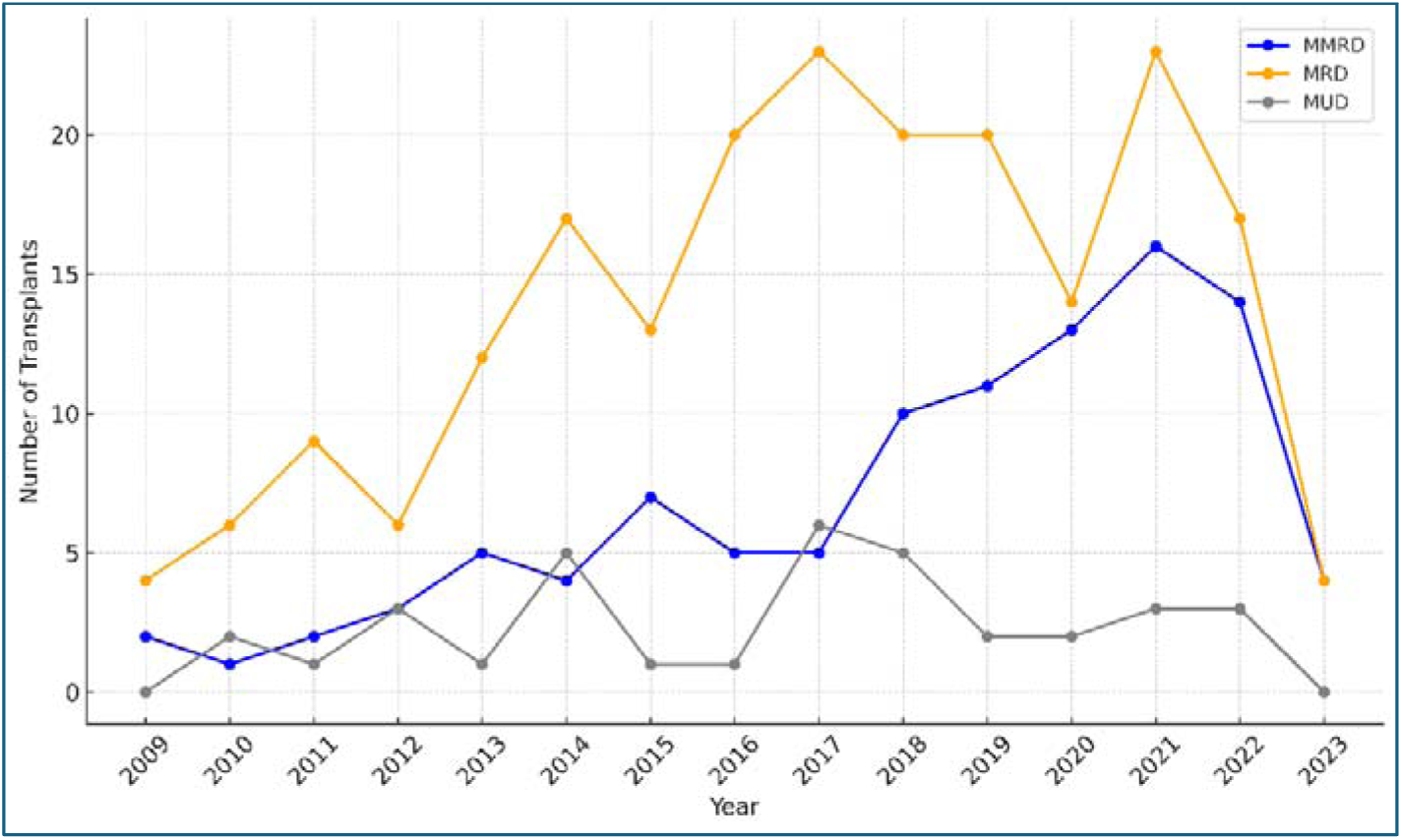
Trends in Donor Types for Allogenic Stem Cell Transplants (2009-2023) *A*nnual trends in allogenic stem cell transplants categorized by donor type from 2009 to 2023. It highlights the fluctuations and trends over the years in the use of Matched Related Donors (MRD), Mismatched Related Donors (MMRD), and Matched Unrelated Donors (MUD).

#### 3.1.4 Nationality and regional distribution

As illustrated in **Supplemental Figure 2**, the majority of transplant recipients were UAE nationals, representing nearly two-thirds of all. Smaller proportions were contributed by patients from the rest of middle east, south Asia, Africa, and other regions, reflecting both national predominance and regional diversity in transplant activity.

### 3.2 Overall Survival

#### 3.2.1 Adult recipients

Kaplan–Meier (KM) overall survival (OS) estimates for adult HCT recipients are summarized in **Table 3** and illustrated in **Figure 2a**. In the allogeneic cohort, AML patients (n=54) had 1-, 3-and 5-year OS rates of 96.2%, 87.2%, and 79.8%, respectively, while ALL patients (n=38) achieved median OS rates of 88.4%, 78.0%, and 78.0% at the same time points. In the autologous HCTs, NHL (n=22) demonstrated a 1-year OS of 95.1%, declining to 77.1% at 5 years, whereas PCDs including MM (n=41) showed a 1-year OS of 97.5% and 5-year OS of 76.1%. KM analysis was restricted to disorders with ≥20 evaluable patients, while crude survival for rarer indications was calculated separately and is provided in **Supplemental Table 5 and 6**.

**Figure 2.**
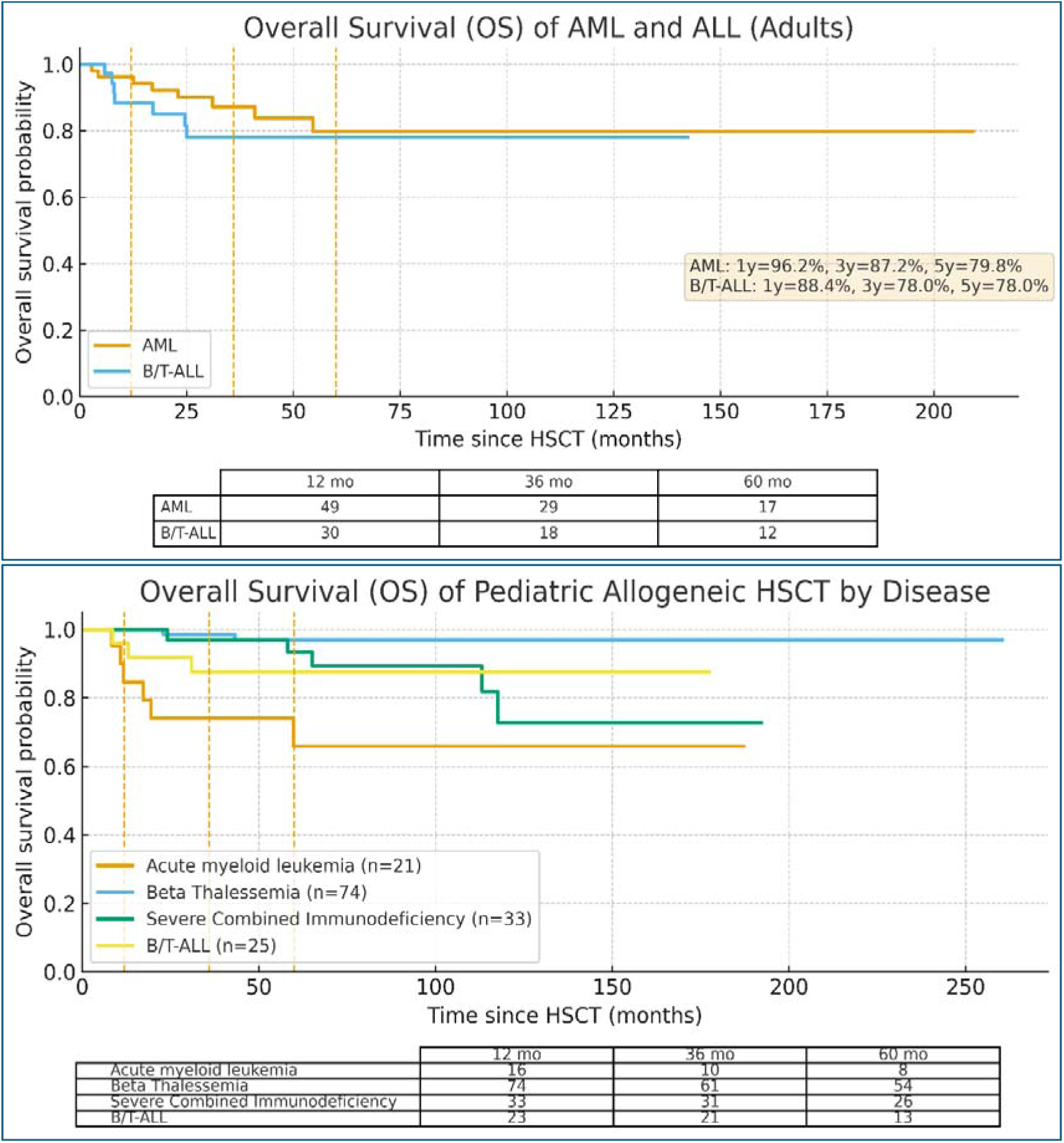
Overall survival (OS) of adult and pediatric patients undergoing allogeneic HCT stratified by underlying disease. Figure 2a. OS of adult patients with acute myeloid leukemia (AML) and B/T-cell acute lymphoblastic leukemia (B/T-ALL) undergoing allogeneic HCT. *KM survival curves show OS at 1, 3, and 5 years, with numbers at risk displayed below the graph*. Figure 2b. OS of pediatric patients undergoing allogeneic HCT stratified by underlying disease. *KM survival curves are summarized for AML,* β *thalassemia, severe combined immunodeficiency (SCID), and B/T-ALL. Numbers at risk are provided below the graph*.

**Table 3.** Overall survival estimates after HCT in adults by disorder.

| Disorder | N | Deaths | 1y OS, %<br>(95% CI) | 3y OS, %<br>(95% CI) | 5y OS, %<br>(95% CI) |
| --- | --- | --- | --- | --- | --- |
| **Allo-HCT** |  |  |  |  |  |
| > AML | 54 | 8 | 96.2 (87.9–100) | 87.2 (77.4–96.9) | 79.8 (66.5–93.1) |
| > ALL | 38 | 7 | 88.4 (77.7–99.1) | 78.0 (63.5–92.6) | 78.0 (63.5–92.6) |
| **Auto-HCT** |  |  |  |  |  |
| > NHL | 22 | 4 | 95.1 (71.9–99.3) | 88.1 (58.8–97.0) | 77.1 (41.2–92.6) |
| > PCD | 41 | 11 | 97.5 (83.5–99.6) | 79.8 (62.0–89.9) | 76.1 (57.6–87.4) |

**Notes:**

#### 3.2.2 Pediatric recipients

KM survival estimates for pediatric HCT recipients are summarized in **Table 4** and illustrated in **Figure 2b**. Among malignant indications, AML (n=21) had 1-, 3-, and 5-year OS rates of 84.7%, 74.1%, and 65.8%, respectively, while ALL (n=25) achieved superior outcomes with OS of 96.0%, 87.7%, and 87.7% at the same landmarks. In the non-malignant group, β-thalassemia (n=74) demonstrated excellent long-term survival with 1-, 3-, and 5-year OS of 100%, 98.6%, and 96.9%, respectively. Similarly, children with SCID (n=33) achieved a 1-year OS of 100%, with sustained survival of 97.0% at 3 years and 93.4% at 5 years. KM analysis was limited to disorders with ≥20 patients, while crude survival proportions for all other indications are detailed in **Supplemental Table 7.**

**Table 4.** Overall survival estimates after HCT in pediatric patients, by disorder.

| Disorder | N | Deaths | 1y OS(95% CI) | 3y OS(95% CI) | 5y OS(95% CI) |
| --- | --- | --- | --- | --- | --- |
| AML | 21 | 6 | 84.7 (59.5-94.8) | 74.1 (48.4-88.3) | 65.8 (38.0-83.5) |
| ALL | 25 | 3 | 96 (74.8-99.4) | 87.7 (66.4-95.8) | 87.7 (66.4-95.8) |
| β-Thalassemia | 74 | 2 | 100 | 98.6 (90.3-99.8) | 96.9 (88.2-99.2) |
| SCID | 33 | 5 | 100 | 97.0 (80.4-99.6) | 93.4 (75.9-98.3) |

- OS = time from HCT to death; survivors/LTFU censored at last contact. N = evaluable patients; Deaths = events by data cut-off. CI = confidence interval. - Survival estimates are based on KM analysis, with curves calculated only for disorders with $\geq 20$ patients, while crude survival was reported for rarer diseases in the Supplementary Table.

In the pediatric cohort undergoing autologous HCT (crude survival outcomes reported in **Supplemental Table 8**), neuroblastoma (n=9), 1-year overall survival (OS) was 88.9%, declined to 44.4% in 3 years and 33.3% at 5 years, with notable loss to follow-up and deaths beyond the first year. In contrast, outcomes for HL(n=3) and medulloblastoma(n=3) appeared more favorable, with 1-year OS of 100% in both groups and 3- and 5-year OS of 66.7%. Given the small numbers in each subgroup, no formal comparisons between diagnoses were attempted.

### 3.3 GVHD incidence and survival impact

#### 3.3.1 Overall incidence and impact on survival

The cumulative incidence of GVHD and its impact on overall survival differed between adults and children is given in **Table 5**. Among adults, the incidence of any GVHD was 69.2% (95% CI, 60.9–76.4), with acute and chronic GVHD occurring in 36.8% (95% CI, 29.1–45.3) and 48.9% (95% CI, 40.5–57.3), respectively. There were no statistically significant differences in OS between patients with and without GVHD in any category (p >0.05). In contrast, pediatric patients showed lower overall GVHD incidence at 45.7% (95% CI, 39.3–52.1), with acute and chronic GVHD affecting 27.8% (95% CI, 22.4–33.9) and 30.4% (95% CI, 24.9–36.7), respectively. Notably, chronic GVHD in children was associated with significantly inferior OS compared with those without chronic GVHD (p=0.023), whereas acute GVHD and overall GVHD did not significantly influence OS.

**Table 5.** I**ncidence and OS according to GVHD type in adults**

| GVHD type | Incidence %<br>(95% CI) | n with<br>GVHD | Deaths | n without<br>GVHD | Deaths | *p* |
| --- | --- | --- | --- | --- | --- | --- |
| **Adults** |  |  |  |  |  |  |
| > **Any type** | 69.2 (60.9-76.4) | 92 | 16 | 41 | 2 | 0.832 |
| > **Acute** | 36.8 (29.1-45.3) | 49 | 9 | 84 | 9 | 0.198 |
| > **Chronic** | 48.9 (40.5-57.3) | 18 | 5 | 52 | 13 | 0.731 |
| **Children** |  |  |  |  |  |  |
| > **Any type** | 45.7 (39.3-52.1) | 105 | 11 | 125 | 8 | 0.234 |
| > **Acute** | 27.8 (22.4-33.9) | 64 | 7 | 166 | 12 | 0.358 |
| > **Chronic** | 30.4 (24.9-36.7) | 70 | 10 | 160 | 9 | 0.023 |

Note: OS = overall survival. *p* values derived from log-rank tests comparing GVHD versus no GVHD for each category.

#### 3.3.2 Adults: Incidence by disease group

Among adult AML recipients (n=54), GVHD occurred in 37 patients (68.5%), including acute GVHD in 19 (35.2%), chronic GVHD in 25 (46.3%), and both in 7 (13.0%). Similarly, in ALL patients (n=38), GVHD developed in 27 cases (71.1%), with acute GVHD in 15 (39.5%), chronic GVHD in 16 (42.1%), and both in 4(10.5%). These findings demonstrate a high burden of GVHD in both myeloid and lymphoid malignancies **Table 6**. For detailed GVHD incidence across all individual disease categories, including rare indications, please refer to **Supplemental Table 9**.

**Table 6.** Incidence of GVHD after allogeneic HCT in adults, by disorder.

| Disorder | N | Any GVHD | aGVHD | cGVHD | Both a+c | No GVHD |
| --- | --- | --- | --- | --- | --- | --- |
| **AML** | 54 | 37 (68.5) | 19 (35.2) | 25 (46.3) | 7 (13.0) | 17 (31.5) |
| **ALL** | 38 | 27 (71.1) | 15 (39.5) | 16 (42.1) | 4 (10.5) | 11 (28.9) |

Notes: aGVHD = acute GVHD (<100 days); cGVHD = chronic GVHD (≥100 days). “Any GVHD” includes patients with aGVHD and/or cGVHD; “Only” categories are mutually exclusive; “Both” indicates patients who developed both acute and chronic GVHD. Percentages are of row totals and may not sum to 100 due to rounding.

#### 3.3.3 Adults: Survival impact by disease group

In AML patients, the presence of acute GVHD was associated with inferior outcomes, with a 5-year OS of 67.9% compared to 86.2% in those without acute GVHD (*p=0.036*). In contrast, the development of chronic GVHD did not significantly affect survival, with comparable 5-year OS of 81.3% versus 79.0% in patients with and without chronic GVHD (*p=0.48*). In ALL patients, survival tended to be lower in those with acute or chronic GVHD, although differences were not statistically significant: 5-year OS was 71.1% versus 83.2% for acute GVHD (*p=0.35*) and 72.5% versus 83.5% for chronic GVHD (*p=0.50*) **Table 7**. These findings suggest that while acute GVHD adversely impacts survival in AML, the effect is less pronounced in ALL, and chronic GVHD does not appear to confer a survival advantage in either group.

**Table 7.**
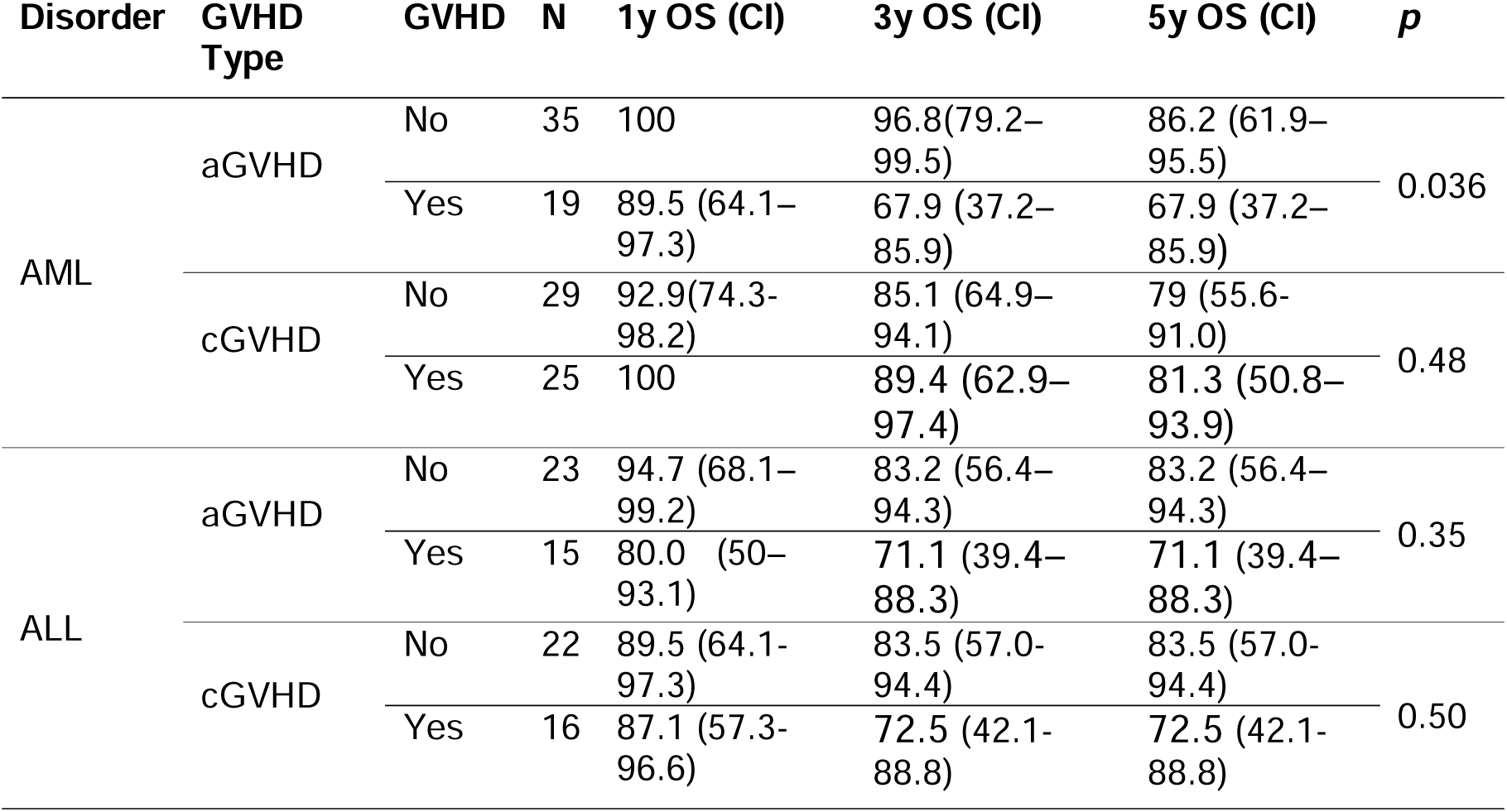
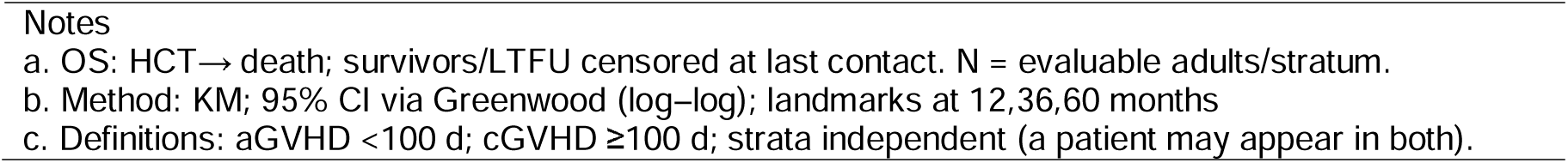
Survival estimates for AML and ALL adult patients stratified by acute and chronic GVHD.

| Disorder | GVHD Type | GVHD | N | 1y OS (CI) | 3y OS (CI) | 5y OS (CI) | p |
| --- | --- | --- | --- | --- | --- | --- | --- |
| AML | aGVHD | No | 35 | 100 | 96.8(79.2–99.5) | 86.2 (61.9–95.5) | 0.036 |
|  |  | Yes | 19 | 89.5 (64.1–97.3) | 67.9 (37.2–85.9) | 67.9 (37.2–85.9) |  |
|  | cGVHD | No | 29 | 92.9(74.3–98.2) | 85.1 (64.9–94.1) | 79 (55.6–91.0) | 0.48 |
|  |  | Yes | 25 | 100 | 89.4 (62.9–97.4) | 81.3 (50.8–93.9) |  |
| ALL | aGVHD | No | 23 | 94.7 (68.1–99.2) | 83.2 (56.4–94.3) | 83.2 (56.4–94.3) | 0.35 |
|  |  | Yes | 15 | 80.0 (50–93.1) | 71.1 (39.4–88.3) | 71.1 (39.4–88.3) |  |
|  | cGVHD | No | 22 | 89.5 (64.1–97.3) | 83.5 (57.0–94.4) | 83.5 (57.0–94.4) | 0.50 |
|  |  | Yes | 16 | 87.1 (57.3–96.6) | 72.5 (42.1–88.8) | 72.5 (42.1–88.8) |  |

#### 3.3.4 Pediatric: Incidence by disease group

The incidence of graft-versus-host disease (GVHD) following allogeneic HCT in pediatric patients is summarized in **Table 8**. Among AML recipients (n=21), GVHD occurred in 11 patients (52.4%), including acute GVHD in 8 (38.1%), chronic GVHD in 9 (42.8%), and both forms in 6 (28.6%). In ALL patients (n=25), GVHD developed in 19 cases (76.0%), with acute GVHD in 14 (56.0%), chronic GVHD in 12 (48.0%), and both in 5 (20.0%). In the non-malignant setting, GVHD was reported in 27 (36.5%) of 74 β-thalassemia patients and 12 (36.4%) of 33 SCID patients, with most cases presenting as chronic or combined GVHD. These findings suggest that GVHD is more frequent in pediatric lymphoid malignancies compared with myeloid or non-malignant disorders. For detailed GVHD incidence across all individual pediatric disease categories, please refer to **Supplemental Table 10**.

**Table 8.** Incidence of GVHD after allogeneic HCT in pediatric patients, by disorder.

| Disorder | N | Any GVHD<br>n(%) | aGVHD,<br>n(%) | cGVHD,<br>n(%) | Both a+c,<br>n(%) | No GVHD,<br>n(%) |
| --- | --- | --- | --- | --- | --- | --- |
| **AML** | 21 | 11 (52.4) | 08 (38.1) | 09 (42.8) | 6 (28.6) | 10 (47.6) |
| **ALL** | 25 | 19 (76.0) | 14 (56.0) | 12 (48.0) | 5 (20.0) | 6 (24.0) |
| **β-thalassemia** | 74 | 27 (36.5) | 11 (14.9) | 22 (29.7) | 6 (8.10) | 47 (63.5) |
| **SCID** | 33 | 12 (36.4) | 05 (15.2) | 07 (21.2) | 0 (0.00) | 21 (63.6) |

Notes: aGVHD = acute GVHD (<100 days); cGVHD = chronic GVHD (≥100 days). “Any GVHD” includes aGVHD and/or cGVHD. Row percentages may not sum to exactly 100 due to rounding.

#### 3.3.5 Pediatrics: Survival impact by disease group

KM survival estimates stratified by GVHD status in pediatric patients are summarized in **Table 9**. In AML (n=21), the presence of acute or chronic GVHD was associated with lower 5-year survival, OS of 46.9% and 47.6%, respectively, compared with 100% and 81.8% in patients without GVHD. In ALL (n=25), survival remained favorable regardless of GVHD status, with 5-year OS ranging from 83.3% to 92.3%, and no significant differences by acute or chronic GVHD. In contrast, outcomes for non-malignant disorders were excellent overall. In β-thalassemia (n=74), 5-year OS exceeded 90% in both acute and chronic GVHD groups, while SCID (n=33) showed similarly high survival, with all patients alive at 5 years except for a modest decline in those with chronic GVHD (85.7%). These results indicate that, unlike in adult AML, the presence of GVHD had limited impact on survival in pediatric ALL and non- malignant disorders, though pediatric AML patients with GVHD demonstrated poorer long-term outcomes.

**Table 9.** Kaplan–Meier survival estimates for AML and ALL, Β-Thalassemia, SCID pediatric patients stratified by acute and chronic GVHD.

|  | GVHD Type | GVHD | N | 1y OS (95% CI) | 3y OS (95% CI) | 5y OS (95% CI) | p |
| --- | --- | --- | --- | --- | --- | --- | --- |
| **AML** | aGVHD | No | 13 | 100 | 100 | 100 | 0.18 |
|  |  | Yes | 8 | 75.0 (31.5-93.1) | 62.5 (22.9-86.1) | 46.9 (12.0-76.3) |  |
|  | cGVHD | No | 12 | 90.9 (50.8-98.7) | 81.8 (44.7-95.1) | 81.8 (44.7-95.1) | 0.19 |
|  |  | Yes | 9 | 76.2 (33.2-93.5) | 63.5 (23.8-86.6) | 47.6 (12.3-76.9) |  |
| **ALL** | aGVHD | No | 12 | 91.7 (53.9-98.8) | 91.7 (53.9-98.8) | 91.7 (53.9-98.8) | 0.59 |
|  |  | Yes | 13 | 100 | 83.3 (48.2-95.6) | 83.3 (48.2-95.6) |  |
|  | cGVHD | No | 14 | 100 | 92.3 (56.6-98.9) | 92.3 (56.6-98.9) | 0.43 |
|  |  | Yes | 11 | 90.9 (50.8-98.7) | 81.8 (44.7-95.1) | 81.8 (44.7-95.1) |  |
| **β-Thalassemia** | aGVHD | No | 63 | 100 | 100 | 98.1 (87.1-99.7) | 0.15 |
|  |  | Yes | 11 | 100 | 90.9 (50.8-98.7) | 90.9 (50.8-98.7) |  |
|  | cGVHD | No | 52 | 100 | 100 | 97.7 (84.6-99.7) | 0.51 |
|  |  | Yes | 22 | 100 | 95.2 (70.7-99.3) | 95.2 (70.7-99.3) |  |
| **SCID** | aGVHD | No | 28 | 100 | 96.4 (77.2-99.5) | 96.4 (77.2-99.5) | 0.34 |
|  |  | Yes | 5 | 100 | 100 | 100 |  |
|  | cGVHD | No | 26 | 100 | 100 | 100 | 0.29 |
|  |  | Yes | 7 | 100 | 85.7 (33.4-97.9) | 85.7 (33.4-97.9) |  |

**Notes**
a. OS: HCT→ death; survivors/LTFU censored at last contact. N = evaluable pediatric HCT per stratum.
b. Method: Kaplan–Meier; 95% CI; landmarks at 12,36,60 months
c. Definitions: aGVHD <100 d; cGVHD ≥100 d. aGVHD and cGVHD are separate strata; a patient may appear in both.

## 4 Discussion

Until 2022, most patients in the UAE requiring HCT were obliged to travel abroad for the procedure. Since then, domestic transplant services have become available, though rarely do some patients still prefer to undergo HCT overseas, often influenced by word-of-mouth experiences from earlier recipients[11]. Despite the high volume of patients treated abroad, there has been a paucity of systematic data describing their outcomes. We previously reported a single-center experience focusing on indications, baseline characteristics, and GVHD incidence, which suggested a relatively high burden of GVHD[12]. Building on that work, the present study provides the first comprehensive report of 454 patients who underwent HCT abroad and were subsequently followed at all three tertiary centers in Abu Dhabi. In this report, we highlight the major findings in the main text, while detailed diagnosis-level distributions, crude survival calculations, and GVHD incidence by disease category are provided in the **supplemental data**.

In adults, AML and ALL were the leading malignant indications for allogeneic HCT, while plasma cell disorders and lymphomas accounted for the majority of autologous HCTs, findings that are consistent with published literature[1][4]. In most pediatric transplant series, malignancies predominate; however, in our cohort non-malignant disorders, particularly thalassemia and primary immune deficiencies, formed the majority, reflecting the higher prevalence of these conditions in the Gulf region[13][14][15] (**Table 1).**

Adults with AML in our cohort underwent allogeneic HCT at a younger median age (45.5 years) compared to Western series, where median ages typically range between 55–65 years, reflecting referral practices that favor younger transplant candidates[16][17]. A male predominance was observed among adults with myeloid and lymphoid malignancies, consistent with published literature[18]. In contrast, within our pediatric cohort (**Table 2**, **Supplemental Table 1**), females were more frequent not only in myeloid and lymphoid malignancies but also in SCID and SCD. This finding contrasts with most published reports, which generally describe a male predominance in these disorders[19]. One possible explanation for this discrepancy may be higher early mortality in male patients, preventing them from reaching transplant eligibility, a phenomenon suggested in survival analyses of both SCID and SCD[20].

In our cohort, the median age of adults undergoing autologous HCT for lymphoid malignancies was 50.1 years[21]. Among pediatric patients, solid tumors represented the predominant indication, with a median age of 3.7 years. These patterns of age distribution and disease indications closely align with prior reports, underscoring the consistency of our findings with the broader autologous HCT reports[22]

Overall, an increasing trend in the number of HCTs performed was observed across the study period. MRD transplants constitute the predominant donor type, followed by MMRD transplants[23][1]. This pattern of raising MMRD reflects the global expansion of haploidentical HCT, largely attributable to improved outcomes achieved through effective GVHD prophylaxis strategies[24]. In contrast, the use of MUD was minimal across both cohorts, which stands in marked contrast to the trends reported by CIBMTR, likely reflecting the challenges of transplant tourism and lack of donor registries. A transient decline in transplant activity was noted during the COVID-19 pandemic[1].

In our cohort, 5-year OS for AML and ALL after allo-HCT was 79.8% and 78.0%,respectively **Table 3**. While these estimates appear higher than that reported in Western series, the wide confidence intervals, small sample size, and few events limit their statistical significance, making them indicative rather than definitive. Crude survival was considerably lower (31.5%; **Supplemental Table 4**), reflecting the impact of follow-up loss, which inflates KM estimates. Different survival end points commonly reported are TRM, disease free survival(DFS) but some studies reported survival using censored quantile regression which reported 5-year OS of 58% [25][26]. When mitigating both KM and crude survival in our report, our mortality outcomes appear broadly consistent with the studies that used OS. However, incomplete data from transplants performed abroad and high loss to follow-up restricted accurate reporting of TRM, DFS, and leukemia-specific mortality.

In adults undergoing autologous HCT, 5-year OS was 77.1% for NHL and 76.1% for plasma-cell disorders/multiple myeloma. Point estimates for NHL appear higher than many historical series of NHL treated with autologous HCT, where 5-year OS commonly approximates 50–60% [27]. For PCD/MM, the 5-year OS aligns with contemporary outcomes, where autologous HCT remains standard and long-term survival frequently reaches the 70–80% range[28]. Overall, these data are directionally consistent with international experience. For the remaining disease categories treated with allogenic HCT, crude survival estimates were calculated and are presented in **Supplemental Table 5** and for autologous HCT in **Supplemental Table 6**.

Among pediatric recipients of allogeneic HCT, outcomes differed by underlying diagnosis (**Table 4**). Children with ALL achieved excellent survival, with OS of 87.7%; however, crude survival dropped to 41.1%, underscoring the effect of loss to follow-up [29]. AML showed lower outcomes than ALL, with OS ranging from 65.8% to 84.7%, slightly above some published series, yet crude survival declined to 38%, below reported benchmarks[30]. In contrast, non-malignant disorders demonstrated consistently favorable outcomes, with both β-thalassemia and SCID maintaining survival above 90%, in line with contemporary reports[31][32]. Crude survival outcomes for individual disease indications are detailed in **Supplemental Table 7.**

Among pediatric autologous HCT recipients with neuroblastoma, 1-year crude OS was 88.9%, consistent with published series; however, survival declined to 44.4% at 3 years and 33.3% at 5 years, reflecting the well-recognized high relapse risk in this population[33].

Outcomes for other indications were based on very small numbers and are therefore reported descriptively in **Supplemental Table 8.**

Our observed GVHD incidence patterns diverge from recent CIBMTR reports. The cumulative incidence of any GVHD in adults (69.2%) and pediatric (45.7%)in our series aligns with the upper range of international experience [1]. Pediatric population showed acute GVHD (27.8%) chronic GVHD (30.4%) in the higher end of range[1][34]. Importantly, only chronic GVHD in children translated into a survival disadvantage (p=0.023), a finding that maybe less consistently emphasized in registry data where both acute and chronic GVHD are often linked to adverse outcomes[34]. Since all patients in this study underwent transplantation abroad, it is possible that children who did not develop acute GVHD and experienced early mortality before follow-up in our center were underrepresented, which may partly account for the observed differences compared to CIBMTR cohorts.

In our adult cohort, the incidence of GVHD among patients surviving beyond the early post-transplant phase was substantial, affecting more than two-thirds of AML and ALL recipients (68.5% and 71.1%, respectively;**Table 6**). Acute GVHD occurred in approximately one-third of patients (35.2% in AML, 39.5% in ALL), while chronic GVHD was observed in over 40% (46.3% and 42.1%, respectively; **Table 6**). These rates lie at the higher end of those reported in EBMT and CIBMTR series, where grade II–IV aGVHD occurs in 20–40% and chronic GVHD in 30–50% of adult allograft recipients [1][35]. Survival analyses showed that acute GVHD in AML was associated with significantly inferior long-term outcomes, consistent with its established role in driving early non-relapse mortality[36], whereas chronic GVHD did not significantly compromise survival, mirroring the complex balance of reduced relapse risk and late morbidity described in prior reports[37] (**Table 7**). In ALL, both acute and chronic GVHD were associated with numerically lower survival, though these differences did not reach statistical significance, likely reflecting sample size limitations. Taking together, these findings reaffirm that acute GVHD remains the principal determinant of adverse survival in adult allogenic HCT, while the impact of chronic GVHD is less definitive[38].

In our pediatric cohort, the burden of GVHD was highest among children with acute leukemia, affecting over half of AML patients and nearly three-quarters of those with ALL[39][40]. In contrast, non-malignant conditions such as β-thalassemia and SCID showed substantially lower rates, with chronic GVHD more common than acute[41][42]. Survival analyses mirrored these patterns: in AML, acute GVHD was associated with markedly inferior long-term outcomes, with 5-year OS below 50% compared to full survival in those without, while chronic GVHD also trended toward reduced survival (**Table 9**). In ALL, both acute and chronic GVHD were linked to numerically lower survival, though small numbers limited statistical power. Children with β-thalassemia and SCID maintained excellent survival irrespective of GVHD status, with 5-year OS consistently above 90%. Collectively, these findings emphasize that acute GVHD remains the dominant driver of adverse survival in pediatric AML, whereas chronic GVHD has a more variable influence, and its clinical impact appears attenuated in non-malignant disorders where relapse risk is not a factor.

## 5 Conclusions

This 14-year, multi-center report of 454 patients followed in the UAE after HCT abroad highlights the distinct features and challenges of the transplant tourism model. In contrast to US and European registries where autologous HCT predominates, our cohort was dominated by allogeneic transplants, reflecting prioritization of curative approaches in patients traveling overseas. Adults undergoing allogeneic HCT were younger than in the western reports, a finding shaped both by referral practices aimed at optimizing outcomes and by the UAE’s demographic structure. An unexpected female predominance was observed in the pediatric population. MUD transplants were rare, underscoring the time-sensitive reliance on MRD before travel and lack of donor registries. The incidence of GVHD was high in both adults and children, at the upper range of international reports, consistent with our prior single-center findings and likely related to variability in prophylaxis and fragmented follow-up. These results underscore the need for sustainable local transplant programs, standardized GVHD prevention strategies such as PTCy, and national registries to optimize outcomes and reduce dependence on overseas care.

## 6 Declaration of generative AI and AI-assisted technologies

During the preparation of this work the author(s) used ChatGPT in order to improve the English language. After using this tool/service, the author(s) reviewed and edited the content as needed and take(s) full responsibility for the content of the published article.

## 7 Funding Statement

This work was supported by the **Department of Health – Abu Dhabi (DOH)**, which also funded the article processing charges (APC) for this publication. The data utilized in this study were originally collected as part of a broader DOH-funded initiative aimed at developing a machine-learning–based prediction model for graft-versus-host disease (GVHD). The dataset was subsequently expanded and analyzed to generate the findings presented in this multi-center outcome report.

## 8 Ethics Statement

This study was approved by the Sheikh Shakhbout Medical City Institutional Review Board, Abu Dhabi, UAE (IRB No.: SSMCREC-444, Date:30-11-2022). De-identified data were retrospectively collected from participating centers, and the requirement for individual consent was waived.

## 9 Author contributions

**NS** and **SH** conceptualized and designed the study. **NS, IA, FAM, AAK, RAH, OTA, MHM, and GEG** collected and analyzed the data. **NS** and **SH** performed the statistical analysis. **NS** and **IA** drafted the initial manuscript. **NS** and **SH** critically revised the manuscript for important intellectual content. All authors contributed to data interpretation, reviewed the final version of the manuscript, and approved it for submission.

## 10 Declaration of interests

The authors have no competing interests to declare that are relevant to the content of this article.

## Data Availability

All data produced in the present study are available upon reasonable request to the authors

## Notes

### Competing Interest Statement

The authors have declared no competing interest.

