## Supplementary document for "Long-term outcomes and GVHD in patients receiving hematopoietic cell transplants abroad: A 14-year UAE multi-center cohort"

### Section 1. Transplant Activity and Demographics

Table 1S. Allogeneic HCT activity and demographics by disease group (2009–2023)

| **Disease** | **Recipients (n)** | | | **Age (IQR)** | **Male n (%)** | **Age (IQR)** | **Male n (%)** |
| --- | --- | --- | --- | --- | --- | --- | --- |
|  | **Adult** | **Ped** | **Total** | **Adult** | | **Ped** | |
| **Myeloid** | **60** | **32** | **92** |  |  |  |  |
| AML | 54 | 21 | 75 | 44.5 (33.1-50.6) | 32 (59.2) | 7.4 (3.0-13.9) | 9 (42.8) |
| CML | 2 | 1 | 3 | 48.2 (36.0-60.3) | 2 (100) | 7.4 (-) | 0 |
| MPN/MDS/BPDCN | 4 | 10 | 14 | 59.0 (20.2-63.1) | 3 (75.0) | 9.1 (7.0-14.2) | 6 (60.0) |
| **Lymphoid** | **52** | **27** | **79** |  |  |  |  |
| B/T-ALL | 38 | 25 | 63 | 30.5 (24.5-39.3) | 23 (60.5) | 6.6 (4.4-12.0) | 10 (40.0) |
| CLL | 1 | 0 | 1 | 55.2 | 1 | 0 | 0 |
| PCD | 4 | 0 | 4 | 41.9 (35.6-56.0) | 4 (100) | 0 | 0 |
| HL | 4 | 0 | 4 | 25.4 (21.8-33.9) | 2 (50.0) | 0 | 0 |
| NHL | 5 | 2 | 7 | 44.1 (18.7-52.4) | 4 (80.0) | 9.9 (2.1-17.6) | 2 (100) |
| **Non-malignant** | **21** | **171** | **192** |  |  |  |  |
| Thalassaemia (α/β) | 4 | 75 | 79 | 31.7 (18.8-37.0) | 2 (50.0) | 6.6 (3.8-10.4) | 32 (42.6) |
| SCD | 8 | 17 | 25 | 27.0 (19.7-33.4) | 4 (50.0) | 10.4 (6.9-13.6) | 12 (70.5) |
| AA | 2 | 14 | 16 | 23.5 | 1 (50.0) | 8.0 (4.5-14.2) | 5 (35.7) |
| FA | 2 | 10 | 12 | 20.65 (19.5-21.8) | 2 (100) | 8.35 (5.4-11.8) | 8 (80.0) |
| SCID | 0 | 33 | 33 | 0 | 0 | 1.1 (0.4–2.1) | 21 (63.6) |
| **Others†** | **5** | **14** | **19** | - | - | - | - |
| **HLH** | **0** | **8** | **8** | 0 | 0 | 4.4 (0.6–8.6) | 8 (100) |
| **Abbreviations:** AML, acute myeloid leukaemia; CML, chronic myeloid leukaemia; MDS, myelodysplastic syndromes; MPN, myeloproliferative neoplasms; BPDCN, blastic plasmacytoid dendritic cell neoplasm; ALL, acute lymphoblastic leukaemia; CLL, chronic lymphocytic leukaemia; PCD, plasma cell disorders; HL, Hodgkin lymphoma; NHL, Non Hodgkin lymphoma; SCD, Sickle cell disease; AA, aplastic anaemia; FA, Fanconi anaemia; SCID, severe combined immunodeficiency; HLH, hemophagocytic lymphohistiocytosis; IQR, interquartile range.  † *Others:* Sideroblastic anemia(1), Erythrogenic porphyria(1),Glanzman thrombasthenia(2),Beta mannosidosis(1), Langerhans cell histiocytosis(1), Chronic granulomatous disease(5), Hyper IgM syndrome (2), common variable immunodeficiency(1), dyskeratosis congenita (2), hypogammaglobulinemia(1), paroxysmal nocturnal hemoglobinuria(1), hereditary spherocytosis(1).  Myeloid includes AML, CML, MDS, MPN, and BPDCN.  For strata with small sample size (n < 10), age is reported as median (range) rather than median (IQR). IQR = interquartile range. | | | | | | | |

Table 2S. Autologous HCT activity and demographics by disease group (2009–2023)

| **Disease** | **Recipients (n)** | | | **Adult: Age (IQR)** | **Adult Male n (%)** | **Pediatric Age**  **(IQR)** | **Pediatric Male n**  **(%)** |
| --- | --- | --- | --- | --- | --- | --- | --- |
|  | **Adult** | **Pediatric** | **Total** |  |  |  |  |
| **Lymphoid** | **74** | **3** | **77** |  |  |  |  |
| PCD | 41 | 0 | 41 | 53.3 (45.9-59.8) | 23 (56.0) | - | - |
| HL | 10 | 3 | 13 | 25.0 (20.3-36.3) | 6 (60.0) | 14.6 (9.7-15.2) | 3 (100) |
| NHL | 22 | 0 | 22 | 47.6 (38.8-56.5) | 15 (68.1) | - | - |
| **Solid tumours** | **0** | **12** | **12** |  |  |  |  |
| Neuroblastoma | 0 | 9 | 9 | - | - | 4.0 (3.2-15.5) | 4 (44.4) |
| Medulloblastoma | 0 | 3 | 3 | - | - | 2.4 (2.3-5.9) | 2 (66.6) |
| **Others** | **3** | **0** | **3** | - | - | - | - |
| **Abbreviations:** IQR, interquartile range; PCD, plasma-cell disorders; NHL, Non-Hodgkin lymphoma; HL, Hodgkin lymphoma.  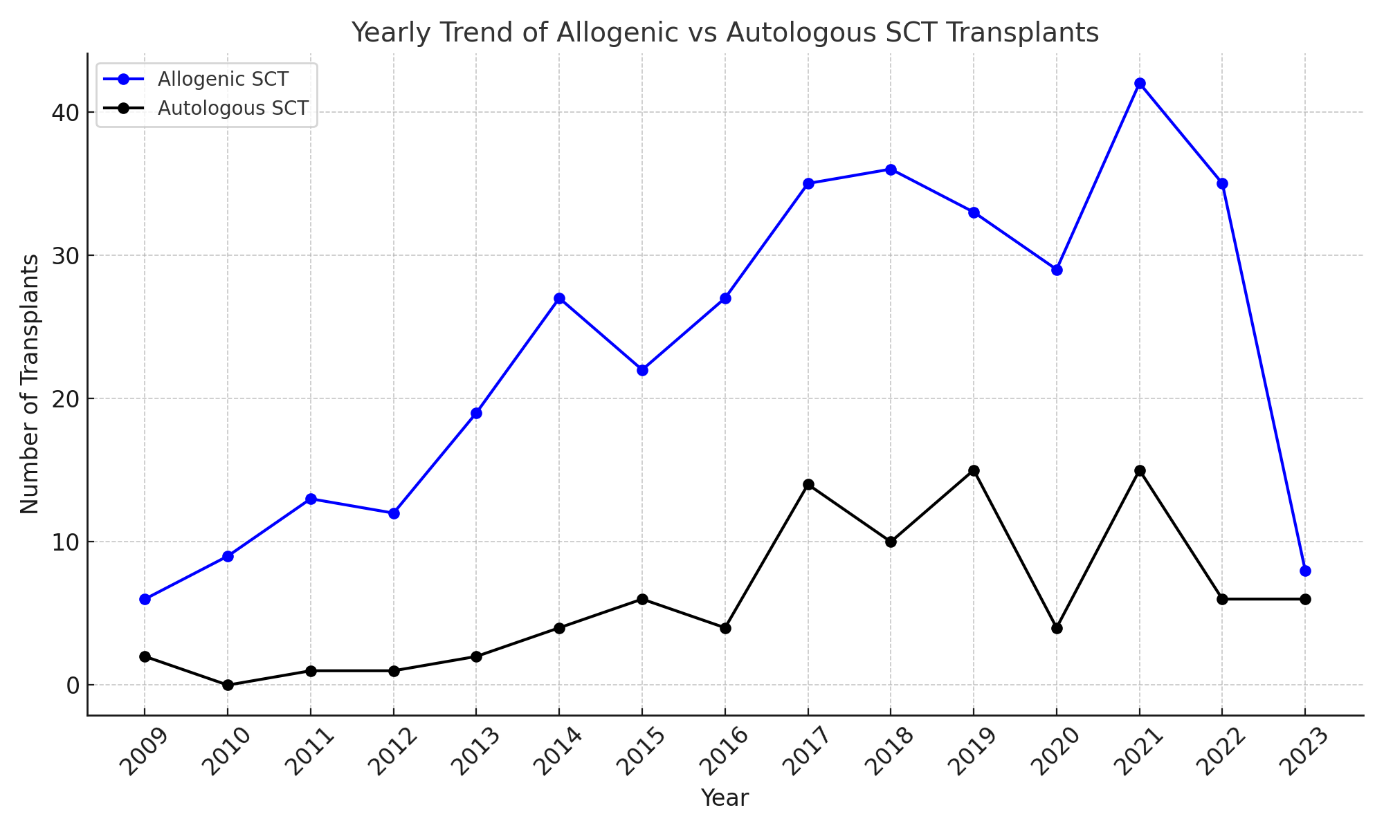  Figure 1S. Yearly Trend of Allogeneic versus Autologous Stem Cell Transplants (2009–2023) Annual number of hematopoietic stem cell transplants performed between 2009 and 2023, stratified into allogeneic (blue line) and autologous (black line) transplants. | | | | | | | |

Table 3S. HCT from 2009 till 2023 by indication, age of recipient at time of transplant, and donor type.

| **Disease Category** | **Allogenic** | | | | | | **Autologous** | | **Total** |
| --- | --- | --- | --- | --- | --- | --- | --- | --- | --- |
|  | **Adult (n)** | | | **Pediatric (n)** | | | **Adult (n)** | **Pediatric (n)** |  |
|  | **HLA-id** | **Haplo ≥ 2MM** | **UK** | **HLA-id** | **Haplo ≥ 2MM** | **UK** |  |  |  |
| **Myeloid** | **46** | **12** | **2** | **19** | **13** | **0** | **1** | **0** | **93** |
| AML | 40 | 12 | 2 | 11 | 9 | 0 | 1 | 0 | 75 |
| CML | 2 | 0 | 0 | 0 | 1 | 0 | 0 | 0 | 3 |
| MPN/MDS/  BPDCN | 4 | 0 | 0 | 7 | 3 | 0 | 0 | 0 | 14 |
| **Lymphoid** | **41** | **10** | **1** | **18** | **9** | **0** | **74** | **3** | **156** |
| B/T-ALL | 28 | 10 | 0 | 17 | 8 | 0 | 1 | 0 | 64 |
| CLL | 1 | 0 | 0 | 0 | 0 | 0 | 0 | 0 | 1 |
| PCD | 4 | 0 | 0 | 0 | 0 | 0 | 41 | 0 | 45 |
| HL | 3 | 0 | 1 | 0 | 0 | 0 | 10 | 3 | 17 |
| NHL | 5 | 0 | 0 | 1 | 1 | 0 | 22 | 0 | 29 |
| **Solid tumors** |  |  |  |  |  |  |  |  |  |
| Neuroblastoma | 0 | 0 | 0 | 0 | 0 | 0 | 0 | 9 | 9 |
| Medulloblastoma | 0 | 0 | 0 | 0 | 0 | 0 | 0 | 3 | 3 |
| **Non-malignant** | **16** | **5** | **0** | **131** | **37** | **3** | **1** | 0 | **193** |
| Thalassemia( α+β) | 3 | 1 | 0 | 53 | 20 | 2 | 0 | 0 | 79 |
| SCD | 7 | 1 | 0 | 13 | 4 | 0 | 0 | 0 | 25 |
| AA | 1 | 1 | 0 | 14 | 0 | 0 | 0 | 0 | 16 |
| FA | 2 | 0 | 0 | 8 | 1 | 1 | 0 | 0 | 12 |
| SCID | 0 | 0 | 0 | 24 | 9 | 0 | 0 | 0 | 33 |
| Other* | 3 | 2 | 0 | 12 | 2 | 0 | 1 | 0 | 20 |
| **HLH** | 0 | 0 | 0 | 7 | 1 | 0 | 0 | 0 | 8 |
| Total patients | **103** | **27** | **3** | **167** | **59** | **3** | **76** | **15** | **454** |
| **Abbreviations:** UK, unknown; AML, acute myeloid leukaemia; CML, chronic myeloid leukaemia; MDS, myelodysplastic syndromes; MPN, myeloproliferative neoplasms; BPDCN, blastic plasmacytoid dendritic cell neoplasm; ALL, acute lymphoblastic leukaemia; CLL, chronic lymphocytic leukaemia; PCD, plasma cell disorders; HL, Hodgkin lymphoma; NHL, Non Hodgkin lymphoma; SCD, Sickle cell disease; AA, aplastic anaemia; FA, Fanconi anaemia; SCID, severe combined immunodeficiency; HLH, hemophagocytic lymphohistiocytosis.  *Sideroblastic anemia(1), Amyloidosis(1), Erythrogenic porphyria(1),Glanzman thrombasthenia(2),Beta mannosidosis(1), Langerhans cell histiocytosis(1), Chronic granulomatous disease(5), Hyper IgM syndrome (2), common variable immunodeficiency(1), dyskeratosis congenita (2), hypogammaglobulinemia(1), paroxysmal nocturnal hemoglobinuria(1), hereditary spherocytosis(1)  The bold text indicates the subtotals of the data listed above in standard text until the next subtotal | | | | | | | | | |

Table 4S. Distribution of donor types by age Group

| Donor type | Adult, n(%) | Pediatric, n(%) |
| --- | --- | --- |
| MRD | 88 (66.1) | 123 (53.8) |
| MMRD | 28 (21.0) | 76 (33.0) |
| MUD | 12 (15.8) | 23 (10.0) |
| MMUD | 0 | 02 (00.9) |
|  | **128** | **114** |
| Percentages are calculated out of the total number of patients in each group.  Abbreviations: MRD, matched related donor; MMRD, mismatched related donor; MUD, matched unrelated donor; MMUD, mismatched unrelated donor. | | |

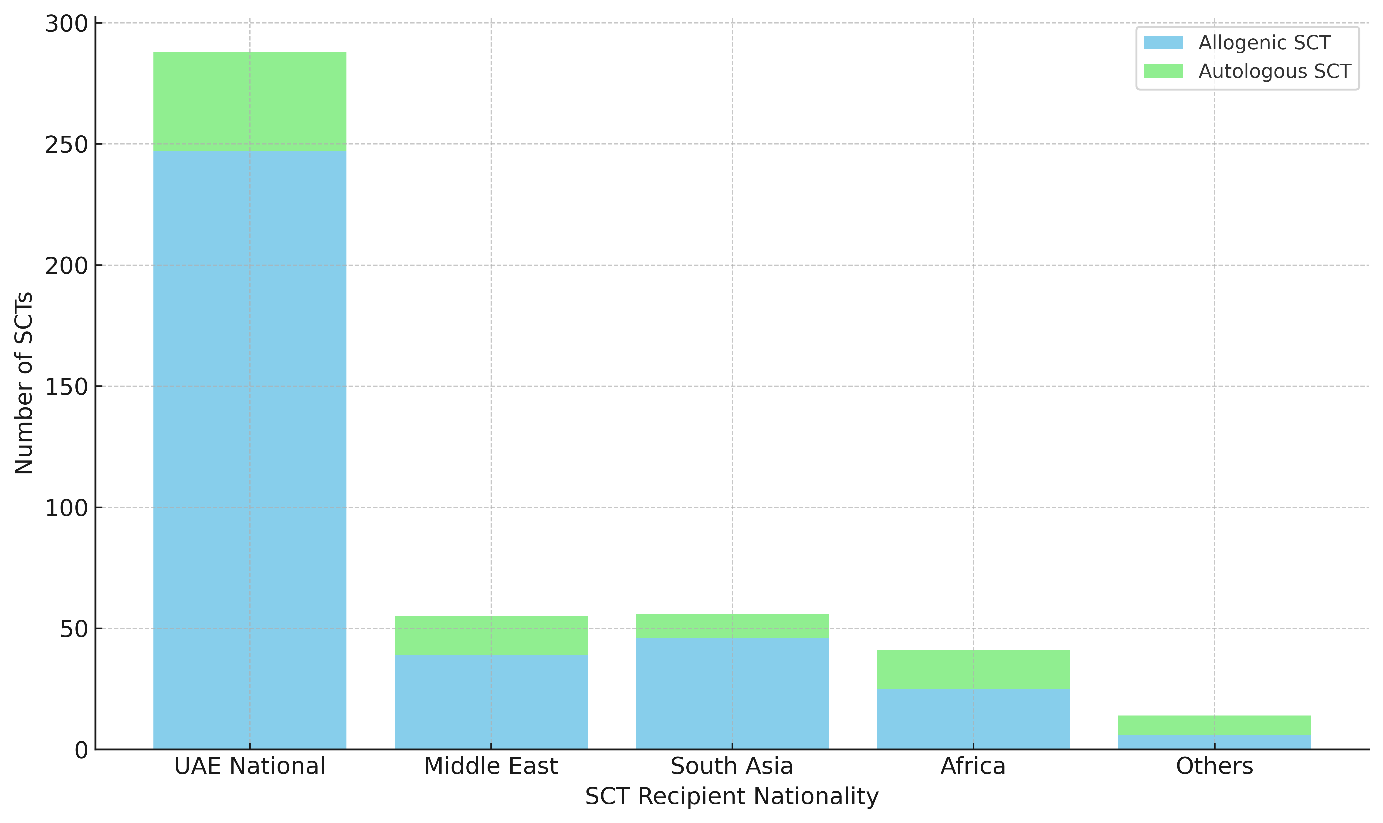

**Figure 2S.** **Allogenic and autologous HCT by recipient nationality.** Distribution of stem cell transplants among different nationalities, with separate stacks indicating the number of allogenic and autologous transplants.

### Section 2. Survival Outcomes

**Table 5S.** **Allogenic HCT crude survival in adults**

|  |  |  | **1 Y** | | |  | **3Y** | | |  | **5Y** | | |
| --- | --- | --- | --- | --- | --- | --- | --- | --- | --- | --- | --- | --- | --- |
|  | **N** |  | **OS** | **LTFU** | **Deaths** |  | **OS** | **LTFU** | **Deaths** |  | **OS** | **LTFU** | **Deaths** |
| AML | 54 |  | 49/54 (90.7) | 3 (5.6) | 2 (3.7) |  | 25/54 (46.3) | 25 (46.3) | 4 (7.4) |  | 17/54 (31.5) | 30 (55.6) | 7 (13.0) |
| ALL | 38 |  | 30/38 (79.0) | 4 (10.5) | 4 (10.5) |  | 18/38 (47.3) | 14 (36.8) | 6 (15.7) |  | 12/38 (31.5) | 20 (52.6) | 6 (15.7) |
| SCD | 8 |  | 8/8 (100) | 0 | 0 |  | 7/8 (87.5) | 1 (12.5) | 0 |  | 6/8 (75.0)* | 2 (25.0) | 0 |
| NHL | 5 |  | 5/5 (100) | 0 | 0 |  | 4/5 (80.0) | 1 (20.0) | 0 |  | 2/5 (20.0) | 3 (60.0) | 0 |
| β-Thal | 4 |  | 4/4 (100) | 0 | 0 |  | 1/4 (25.0) | 3 (75.0) | 0 |  | 0/4 (0) | 4 (100) | 0 |
| HL | 4 |  | 4/4 (100) | 0 | 0 |  | 3/4 (75.0) | 1 (25.0) | 0 |  | 2/4 (50.0) | 2 (50.0) | 0 |
| MDS | 3 |  | 2/3 (66.7) | 0 | 1 (33.3) |  | 1/3 (33.3) | 1 (33.3) | 1 (33.3) |  | 1/3 (33.3) | 1 (33.3) | 1 (33.3) |
| MM | 4 |  | 4/4 (100) | 0 | 0 |  | 3/4 (75.0) | 1 (25.0) | 0 |  | 1/4 (25.0) | 3 (75.0) | 0 |
| AA | 2 |  | 2/2 (100) | 0 | 0 |  | 1/2 (50.0) | 1 (50.0) | 0 |  | 1/2 (50.0) | 1 (50.0) | 0 |
| BPDCN | 1 |  | 1/1 (100) | 0 | 0 |  | 1/1 (100) | 0 | 0 |  | 1/1 (100) | 0 | 0 |
| CGD | 1 |  | 1/1 (100) | 0 | 0 |  | 0/1 (0) | 1 (100) | 0 |  | 0/1 (0) | 1 (100) | 0 |
| CLL | 1 |  | 1/1 (100) | 0 | 0 |  | 1/1 (100) | 0 | 0 |  | 1/1 (100) | 0 | 0 |
| CML | 2 |  | 2/2 (100) | 0 | 0 |  | 2/2 (100) | 0 | 0 |  | 1/2 (50.0) | 1 (50.0) | 0 |
| FA | 2 |  | 2/2 (100) | 0 | 0 |  | 1/2 (50.0) | 1 (50.0) | 0 |  | 0/2 (0) | 2 (100) | 0 |
| GT | 1 |  | 1/1 (100) | 0 | 0 |  | 0/1 (0) | 1 (100) | 0 |  | 0/1 (0) | 1 (100) | 0 |
| HGG | 1 |  | 1/1 (100) | 0 | 0 |  | 1/1 (100) | 0 | 0 |  | 1/1 (100) | 0 | 0 |
| LCH | 1 |  | 1/1 (100) | 0 | 0 |  | 1/1 (100) | 0 | 0 |  | 1/1 (100) | 0 | 0 |
| PNH | 1 |  | 1/1 (100) | 0 | 0 |  | 0/1 (0) | 1 (100) | 0 |  | 0/1 (0) | 1 (100) | 0 |
| - LTFU = lost to follow-up; Crude OS = Crude overall survival. - Crude survival was calculated as the proportion of patients alive at each landmark time point (Alive/Total, %). Survival percentages were calculated excluding patients lost to follow-up and/or with less than 1, 3, or 5 years of follow-up. - *Estimates marked with an asterisk are unreliable due to extremely small patient numbers.*   **Abbreviations:** AML, acute myeloid leukaemia; CML, chronic myeloid leukaemia; MDS, myelodysplastic syndromes; MPN, myeloproliferative neoplasms; BPDCN, blastic plasmacytoid dendritic cell neoplasm; CGD, chronic granulomatous disease; ALL, acute lymphoblastic leukaemia; CLL, chronic lymphocytic leukaemia; PCD, plasma cell disorders; HL, Hodgkin lymphoma; NHL, Non Hodgkin lymphoma; SCD, Sickle cell disease; AA, aplastic anaemia; FA, Fanconi anaemia; SCID, severe combined immunodeficiency; CVID, common variable immunodeficiency; HLH, hemophagocytic lymphohistiocytosis; IQR, interquartile range. | | | | | | | | | | | | | |

**Table 6S. Autologous HCT crude survival rate in adults**

|  |  | **1Y** | | |  | **3Y** | | |  | **5Y** | | |
| --- | --- | --- | --- | --- | --- | --- | --- | --- | --- | --- | --- | --- |
|  | **N** | **OS** | **LTFU** | **Deaths** |  | **OS** | **LTFU** | **Deaths** |  | **OS** | **LTFU** | **Deaths** |
| **PCD** | 41 | 39 (95.1) | 1 (2.4) | 1 (2.4) |  | 25 (61.0) | 11 (26.8) | 5 (12.2) |  | 19 (46.3) | 14 (34.1) | 8 (19.5) |
| **NHL** | 22 | 18 (81.8) | 3 (13.6) | 1 (4.5) |  | 11 (50.0) | 9 (40.9) | 2 (9.1) |  | 7 (31.8) | 12 (54.5) | 3 (13.6) |
| **HL** | 10 | 10 (100) | 0 | 0 |  | 8 (80.0) | 1 (10) | 1 (10.0) |  | 4 (40.0) | 5 (50.0) | 1 (10.0) |
| *Inaccurate estimates due to extremely low number of patients  ^$^Estimated 1/3/5-year survival- removed patients who lost to follow up and/or who have less than 1/3/5 year follow up  **Abbreviations:** LTFU, lost to follow up; PCD, plasma-cell disorders; NHL, Non-Hodgkin lymphoma; HL, Hodgkin lymphoma. | | | | | | | | | | | | |

**Table 7S.** **Allogenic HCT crude survival rate in pediatric patients**

|  |  |  | **1Y** | | |  | **3Y** | | |  | **5Y** | | |
| --- | --- | --- | --- | --- | --- | --- | --- | --- | --- | --- | --- | --- | --- |
| **Disorder** | **N** |  | **OS** | **LTFU** | **Deaths** |  | **OS** | **LTFU** | **Deaths** |  | **OS** | **LTFU** | **Deaths** |
| **β-** **Thalassemia** | 74 |  | 74 (100) | 0 | 0 |  | 61 (82.4) | 12 (16.2) | 1 |  | 54 (73.0) | 18 (24.3) | 2  (2.7) |
| **SCID** | 33 |  | 33 (100) | 0 | 0 |  | 31 (93.9) | 1 (3.0) | 1  (3.0) |  | 26 (78.8) | 5 (15.2) | 2  (6.1) |
| **AML** | 21 |  | 16 (76.2) | 3 (14.3) | 2  (9.5) |  | 10 (47.6) | 6 (28.6) | 5 (23.8) |  | 8 (38.1) | 7 (33.3) | 6 (28.6) |
| **B- ALL** | 19 |  | 17  (89.5) | 1  (5.30) | 1  (5.30) |  | 15  (78.9) | 1  (5.30) | 3  (15.8  ) |  | 8  (42.1) | 8  (42.1) | 3  (15.8) |
| **SCD** | 17 |  | 17  (100) | 0 | 0 |  | 17  (100) | 0 | 0 |  | 14  (82.4)* | 3 (17.6) | 0 |
| **AA** | 14 |  | 12 (85.7) | 2 (14.3) | 0 |  | 9 (64.3) | 5 (35.7) | 0 |  | 8 (57.1) | 6 (42.9) | 0 |
| **FA** | 10 |  | 10 (100) | 0 | 0 |  | 9 (90.0) | 0 | 1 (10.0) |  | 7 (70.0) | 1 (10.0) | 2 (20.0) |
| **HLH** | 8 |  | 7 (87.5) | 1 (12.5) | 0 |  | 4 (50.0) | 4 (50.0) | 0 |  | 4 (50.0) | 4 (50) | 0 |
| **MDS** | 7 |  | 7 (100) | 0 | 0 |  | 6 (85.7) | 0 | 1 (14.3) |  | 5 (71.4) | 1 (14.3) | 1 (14.3) |
| **CGD** | 4 |  | 4 (100) | 0 | 0 |  | 4 (100) | 0 | 0 |  | 1 (25.0) | 3 (75.0) | 0 |
| **α- Thalassemia** | 1 |  | 1 | 0 | 0 |  | 0 | 1 | 0 |  | 0 | 1 | 0 |
| **β- Mannosidosis** | 1 |  | 1 | 0 | 0 |  | 0 | 1 | 0 |  | 0 | 1 | 0 |
| **CML** | 1 |  | 1 | 0 | 0 |  | 0 | 1 | 0 |  | 0 | 1 | 0 |
| **CVID** | 1 |  | 1 | 0 | 0 |  | 1 | 0 | 0 |  | 0 | 1 | 0 |
| **DC** | 2 |  | 2 | 0 | 0 |  | 2 | 0 | 0 |  | 0 | 2 | 0 |
| **GT** | 1 |  | 1 | 0 | 0 |  | 1 | 0 | 0 |  | 1 | 0 | 0 |
| **HS** | 1 |  | 1 | 0 | 0 |  | 1 | 0 | 0 |  | 1 | 0 | 0 |
| **PMF** | 2 |  | 2 | 0 | 0 |  | 2 | 0 | 0 |  | 1 | 1 | 0 |
| **MPN** | 1 |  | 1 | 0 | 0 |  | 1 | 0 | 0 |  | 0 | 1 | 0 |
| **NHL** | 2 |  | 2 | 0 | 0 |  | 1 | 1 | 0 |  | 0 | 2 | 0 |
| **EP** | 1 |  | 1 | 0 | 1 |  | 1 | 0 | 0 |  | 1 | 0 | 0 |
| **SA** | 1 |  | 1 | 0 | 0 |  | 1 | 0 | 0 |  | 1 | 0 | 0 |
| **X- Hyper IgM** | 2 |  | 2 | 0 | 0 |  | 2 | 0 | 0 |  | 2 | 0 | 0 |
| All values are crude survival proportions. Percentages are rounded to one decimal; results for small denominators (n<10) are unstable.  Perfect survival (100%) estimates may reflect small sample size and should be interpreted cautiously.  **Abbreviations:** AA, aplastic anaemia; ALL, acute lymphoblastic leukaemia; AML, acute myeloid leukaemia; BPDCN, blastic plasmacytoid dendritic cell neoplasm; CGD, chronic granulomatous disease; CLL, chronic lymphocytic leukaemia; CML, chronic myeloid leukaemia; CVID, common variable immunodeficiency; FA, Fanconi anaemia; HLH, hemophagocytic lymphohistiocytosis; IQR, interquartile range; MM, multiple myeloma; MDS, myelodysplastic syndromes; MPN, myeloproliferative neoplasms; PCL, plasma-cell leukaemia; GT, Glanzman thrombasthenia; HS, Hereditary spherocytosis; SCID, severe combined immunodeficiency; EP, Erythrogenic porphyria; SA, Sideroblastic anemia. | | | | | | | | | | | | | |

**Table 8S.** **Autologous HSCT crude survival in pediatric patients**

|  |  | **1Y** | | | |  | **3Y** | | | |  | **5Y** | | |
| --- | --- | --- | --- | --- | --- | --- | --- | --- | --- | --- | --- | --- | --- | --- |
| **Disorder** | **N** | **OS** | | **LTFU** | **Deaths** |  | **OS** | | **LTFU** | **Deaths** |  | **OS** | **LTFU** | **Deaths** |
| **Neuroblastoma** | 9 | 8  (88.9) | | 1 (11.1) | 0 |  | 4  (44.4) | | 3 (33.3) | 2 (22.2) |  | 3  (33.3) | 4  (44.4) | 2 (22.2) |
| **HL** | 3 | 3  (100) | | 0 | 0 |  | 2  (66.7) | | 1 (33.3) | 0 |  | 2  (66.7) | 1 (33.3) | 0 |
| **Medulloblastoma** | 3 | 3  (100) | | 0 | 0 |  | 2  (66.7) | | 1 (33.3) | 0 |  | 2  (66.7) | 1 (33.3) | 0 |
| All values are actual (crude) survival proportions at 1/3/5 years after ASCT  *Abbreviations:* HL, Hodgkin lymphoma; OS, overall survival; LTFU, lost to follow-up. | | | | | | | | | | | | | | |

### Section 3. GVHD Incidence

Table 9S. GVHD incidence post allogenic HCT in adults

| **Disorder** | **N** | **GVHD, n%** | **Acute GVHD, n%** | **Chronic GVHD, n%** |
| --- | --- | --- | --- | --- |
| **AML** | 54 | 37 (68.5) | 19 (35.2) | 25 (46.3) |
| **B- ALL** | 31 | 22 (71.0) | 13 (41.9) | 12 (38.7) |
| **T- ALL** | 7 | 5 (71.4) | 2 (28.6) | 4 (57.1) |
| **AA** | 2 | 2 (100) | 1 (50.0) | 1 (50.0) |
| **β- Thalassemia** | 4 | 1 (25.0) | 0 | 1 (25.0) |
| **FA** | 2 | 1 (50.0) | 0 | 1 (50.0) |
| **SCD** | 8 | 3 (37.5) | 1 (12.5) | 3 (37.5) |
| **HL** | 4 | 2 (50.0) | 2 (50) | 1 (25.0) |
| **NHL** | 5 | 5 (100) | 2 (40) | 4 (80.0) |
| **MM** | 4 | 4 (100) | 3 (75) | 3 (75.0) |
| GVHD indicates graft-versus-host disease. Percentages were calculated as the number of affected patients divided by the total number of transplants within each disease category. Acute and chronic GVHD cases are not mutually exclusive, and patients may be counted in both categories.  Acute and chronic GVHD categories are not mutually exclusive  **Abbreviations:** AA, aplastic anaemia; ALL, acute lymphoblastic leukaemia; AML, acute myeloid leukaemia; BPDCN, blastic plasmacytoid dendritic cell neoplasm; CGD, chronic granulomatous disease; CLL, chronic lymphocytic leukaemia; CML, chronic myeloid leukaemia; CVID, common variable immunodeficiency; FA, Fanconi anaemia; HLH, hemophagocytic lymphohistiocytosis; IQR, interquartile range; MM, multiple myeloma; MDS, myelodysplastic syndromes; MPN, myeloproliferative neoplasms; PCL, plasma-cell leukaemia; GT, Glanzman thrombasthenia; HS, Hereditary spherocytosis; SCID, severe combined immunodeficiency; EP, Erythrogenic porphyria; SA, Sideroblastic anemia. | | | | |

Table 10S. GVHD incidence across allogenic HSCT in pediatrics

| **Disorder** | **N** | **GVHD, n %** | **Acute GVHD, n%** | **Chronic GVHD, n%** |
| --- | --- | --- | --- | --- |
| **β- thalassemia** | 74 | 27 (36.5) | 11(14.9) | 22 (29.7) |
| **SCID** | 33 | 12 (36.4) | 5 (15.2) | 7 (21.2) |
| **AML** | 21 | 11 (52.4) | 8 (38.1) | 9 (42.9) |
| **B- ALL** | 19 | 15 (78.9) | 11 (57.9) | 8 (42.1) |
| **SCD** | 17 | 9 (52.9) | 8 (47.1) | 3 (17.6) |
| **AA** | 14 | 6 (42.9) | 5 (35.7) | 3 (21.4) |
| **FA** | 10 | 1 (10) | 1 (10) | 0 |
| **HLH** | 8 | 4 (50) | 3 (37.5) | 2 (25) |
| **MDS** | 7 | 6 (85.7) | 5 (71.4) | 5 (71.4) |
| **T- ALL** | 6 | 4 (66.7) | 2 (33.3) | 3 (50) |
| **NHL** | 2 | 1 | 1 | 0 |
| GVHD indicates graft-versus-host disease. Percentages were calculated as the number of affected patients divided by the total number of transplants within each disease category. Acute and chronic GVHD cases are not mutually exclusive, and patients may be counted in both categories.  **Abbreviations:** SCID, severe combined immunodeficiency; AML, acute myeloid leukaemia; ALL, acute lymphoblastic leukaemia; SCD, Sickle cell disease; AA, aplastic anaemia; FA, Fanconi anaemia; HLH, hemophagocytic lymphohistiocytosis; MDS, myelodysplastic syndromes; NHL, Non-Hodgkin lymphoma. | | | | |
